# Changes in mitral regurgitation after alcohol septal ablation in hypertrophic obstructive cardiomyopathy: a retrospective cohort study

**DOI:** 10.64898/2025.12.03.25341595

**Authors:** Anna Damlin, Andreas Rück, Mesud Mustafic, Rebecka Jandér, Anette Rickenlund, Maria J Eriksson, Nawzad Saleh, David Marlevi

**Affiliations:** Department of Molecular Medicine and Surgery, Division of Clinical Physiology, Karolinska Institutet, Stockholm, Sweden; Department of Cardiology, Karolinska University Hospital, Stockholm, Sweden; Unit of Cardiology, Department of Medicine Solna, Karolinska Institutet, Stockholm, Sweden; Department of Cardiology, Danderyds Hospital, Stockholm, Sweden; Department of Clinical Physiology, Södersjukhuset, Stockholm, Sweden; Department of Clinical Physiology, Karolinska University Hospital, Stockholm, Sweden; Institute for Medical Engineering and Science, Massachusetts Institute of Technology, Cambridge, MA 02139, USA

**Author notes:** **Corresponding author:** Anna Damlin,. K1 Molekylär medicin och kirurgi, 171 76 Stockholm, Sweden.

**Keywords:** Alcohol septal ablation, Echocardiography, Hypertrophic obstructive cardiomyopathy, Mitral regurgitation

## Abstract

**Background:** The primary mechanism of mitral regurgitation (MR) in hypertrophic obstructive cardiomyopathy (HOCM) is systolic anterior motion (SAM) of the mitral leaflets due to septal hypertrophy narrowing the left ventricular outflow tract (LVOT), influencing leaflet movement and closure. Among patients undergoing percutaneous alcohol septal ablation (ASA) to reduce the interventricular septum, the impact on MR and the underlying mechanism of MR reduction remains less studied. Therefore, we aimed to determine the prevalence of MR before and changes in MR severity after ASA, and analyze echocardiographic (ECHO) parameters associated with MR changes.

**Methods:** All patients who underwent ASA at Karolinska University Hospital in Stockholm, Sweden, 2009-2021, were included in this retrospective cohort study, with follow-up until mid 2025. ECHO parameters were collected before ASA, at 1-year follow-up and at the latest follow-up registered. Linear and logistic regression models were used to assess the association between ECHO parameters and the direction of change in MR at follow-up.

**Results:** 154 patients were included with mean follow-up time 4.0±3.9 years. 13.6% (n=21) had at least moderate MR at baseline, of which 90.1% (n=19) decreased and 9.9% (n=2) had persistent moderate MR at the last follow-up. The patients with decreased (n=19, 12.3%) or persistent grade of MR (n=71, 46.1%) showed significant reduction of median basal septal wall thickness and LVOT peak pressure gradients (decreased MR: 20.0mm to 14.0mm, p<0.001, 137.5mmHg to 4.8mmHg, p<0.001, persistent MR: 19.0mm to 16.0mm, p<0.001, 95.0mmHg to 25.0mmHg) between the baseline and the last follow-up ECHO. In contrast, no significant differences were seen in the patients with increased MR grade (n=7, 4.5%, 21.0mm to 18.0mm, p=0.251, 57.0mmHg to 51.0mmHg, p=0.336).

**Conclusion:** In most patients with at least moderate MR prior to ASA, MR decreased at follow-up, as a supposed effect of septal reduction, and reduced SAM.

## Background

The worldwide prevalence of hypertrophic cardiomyopathy is estimated to be 1:200 to 1:500. About 60% to 70% of the patients with hypertrophic cardiomyopathy present with systolic anterior motion (SAM) of the mitral valve (MV) and left ventricular outflow tract (LVOT) obstruction (LVOTO), resting or provoked (inducible), hence defined as hypertrophic obstructive cardiomyopathy (HOCM) (1–3). The pathophysiology behind SAM in HOCM comprises a mitral leaflet-septal contact during systole, together with an abnormal systolic anterior movement of the mitral leaflets towards a hypertrophic interventricular septum, narrowing the LVOT causing LVOTO and impaired mitral valve closure (4, 5). Patients with HOCM are more prone to have structural abnormalities of the MV apparatus including elongated mitral leaflets, papillary muscle malformations, displacement and/or insertion anomalies, and anomalous chordal insertions (6–8), which may contribute to SAM.

Mitral regurgitation (MR) is a common consequence of SAM with or without intrinsic structural abnormalities of the MV apparatus in HOCM (9). The mechanism behind MR in SAM is the anterior displacement of the anterior mitral leaflet in combination with a too short, or stiff posterior mitral leaflet, causing a loss of coaptation (10). Surgical (septal myectomy), or percutaneous (alcohol septal ablation (ASA)) reduction of the interventricular septum aiming to reduce LVOTO often improves SAM and MR in patients with HOCM (11–14). Three studies demonstrate reduction of MR after ASA, however none of the studies had MR as the primary outcome and did not further analyze the mechanism behind MR and factors associated with MR progression or reduction after ASA (15–17). In this study, we aim to describe changes in MR severity after ASA, and to compare the clinical characteristics and echocardiographic parameters in patients who had increased grade of MR with those who had an improvement of pre-existing MR, or persistent grade of MR after ASA.

## Methods

### Study design and population

This retrospective study comprised a population of patients with HOCM that underwent ASA on clinical indication between June 2, 2009, and July 1, 2021, at the Karolinska University Hospital in Stockholm, Sweden, which serves as a tertiary referral center, enabling treatment for patients referred from different regions in Sweden. The study population has been previously described (18).

### Data collection

Patient- as well as ASA specific data were collected from patient records. All patients underwent a pre-procedural transthoracic echocardiography (ECHO) at maximum one week prior to ASA with pre-procedural ECHO parameters collected (listed in the next paragraph) as well as age, sex, length, weight, and body surface area (BSA). All patients did at least one follow-up ECHO during the follow-up period (from ASA until July 1, 2025), which was scheduled at one year after the procedure as by clinical routine. If performed, data from an additional follow-up ECHO (the latest ECHO performed during the follow-up period) was included in the study, i.e. three ECHO examinations per patient were analyzed in total. The ECHO parameters were collected from the imaging system (Viewpoint, GE HealthCare, U.S.A.). Specially trained ECHO technicians or medical doctors specialized in cardiovascular imaging performed the ECHO examinations. The image acquisition was performed according to standard protocol including additional images focusing on HOCM specific morphology and Doppler findings (as described below) to enable comprehensive HOCM evaluation (14, 19, 20). The images were reviewed by at least one medical doctor specialized in cardiovascular imaging before the examination was finished, to enable additional images if needed. All images were retrospectively reviewed by the first author (medical doctor specialized in cardiovascular imaging), not viewing the clinical reports from the ECHO examinations prior to the review. After the study review, the study report was compared with the clinical report, and if any differences occurred, images were reviewed with an additional cardiovascular imaging specialist to reach consensus.

The data collected from the pre-procedural ECHO were heart rate, LV end-diastolic and end-systolic diameters and volumes, LV ejection fraction, E/A, E/é (measurements used in ECHO to assess left ventricular diastolic function), left atrium end-systolic volume, LV basal septal wall thickness and posterior wall thickness (20, 21). Also, the grade of mitral- and aortic regurgitation according to the European Association of Cardiology guidelines (22), presence of SAM and LVOT peak systolic gradients, ΔP, at rest and at Valsalva maneuver, estimated from LVOT velocity (V), using the simplified Bernoulli equation (ΔP=4V^2^) were collected. The length of the MV leaflets and coaptation length were manually measured (20, 23) from the pre-procedural ECHO images. The measurements of the anterior mitral leaflet (AML) and posterior mitral leaflet (PML) length were made in the apical three-chamber view at end-diastole (n=145, 94.2%). Among patients with poor visualization in apical view, the measurements were performed in parasternal long axis view (PLAX) (n=9, 5.8%). At follow-up, ECHO data and information about any repeat ASA or myectomy, as well as mitral valve surgery were collected. Follow-up ECHO variables were the same as from the pre-procedural ECHOs except for LV posterior wall thickness that was not collected from the follow-up ECHOs.

### Statistical analysis

To assess possible normal distribution, a Shapiro-Wilk test was used on collected variables. Normally distributed data were presented with mean ± standard deviation (SD). Non-normally distributed data were presented with median and interquartile range (IQR). Binary variables were reported as absolute numbers and percentages. The patients were grouped depending on if they had an increase in MR grade, persistent grade of MR, or a decrease in MR grade at the last follow-up. Upon review of ECHO images and reports, intermediate grades of MR were categorized as follows: trivial to mild MR was classified as mild MR; mild to moderate as moderate MR; moderate-severe MR as severe MR (22). An increase in MR severity was defined as progression from no MR or mild MR on pre-procedural ECHO to moderate or severe MR on follow-up, or from pre-procedural moderate MR to severe MR on follow-up. A decrease in MR severity was defined as improvement from moderate MR pre-procedure to mild or no MR on follow-up, or from severe MR pre-procedure to moderate, mild, or no MR on follow-up. A change from no pre-procedural MR to mild MR on follow-up was not considered as increased MR. To evaluate the correlation between continuous or binary values and outcome, linear or logistic regression models were executed. As there were quite few cases in the increased/decreased MR groups, no multivariate analyses were performed. P-values <0.05 were considered significant. Data analysis was conducted using STATA software (version 17.0 Stata Corp., College Station, Texas, USA).

## Results

In total, 154 patients with HOCM that underwent ASA were included (Table 1). Half of the patients were women, and the mean age was 61.0 ± 12.0 years. Of the included patients, all had SAM, and 13.6% (n=21) had at least moderate MR at the pre-procedural ECHO (Table 2). The mean follow-up time from ASA to the latest follow-up ECHO was 4.0 ± 3.9 years (median 2.7 years, IQR 1.0-5.7 years). Of the patients with at least moderate MR at the pre-procedural ECHO, 90.1% (n=19) demonstrated improvement to mild MR or less (with no mitral valve intervention done), while 9.9% (n=2) had persistent moderate MR at the last follow-up (Fig. 1). At the last follow-up, 46.1% (n=71) patients had persistent grade of MR (mild grade: n=69, moderate grade: n=2). In total, 4.5% (n=7) patients increased from mild or no pre-procedural MR to moderate MR at follow-up. No patients had severe MR at follow-up. After grouping the patients as increased, persistent, or decreased MR, 57 patients remained ungrouped as they had either no MR at baseline and mild MR at follow-up, or mild MR at baseline and no MR at follow-up. The follow-up time among the patients that had an increased grade of MR was 5.3 ± 4.7 years, compared with 3.1 ± 3.0 years among the patients with decreased MR grade (p=0.168), and 3.9 ± 3.8 years among those with persistent grade of MR at the last follow-up (p=0.376).

**Fig. 1.**
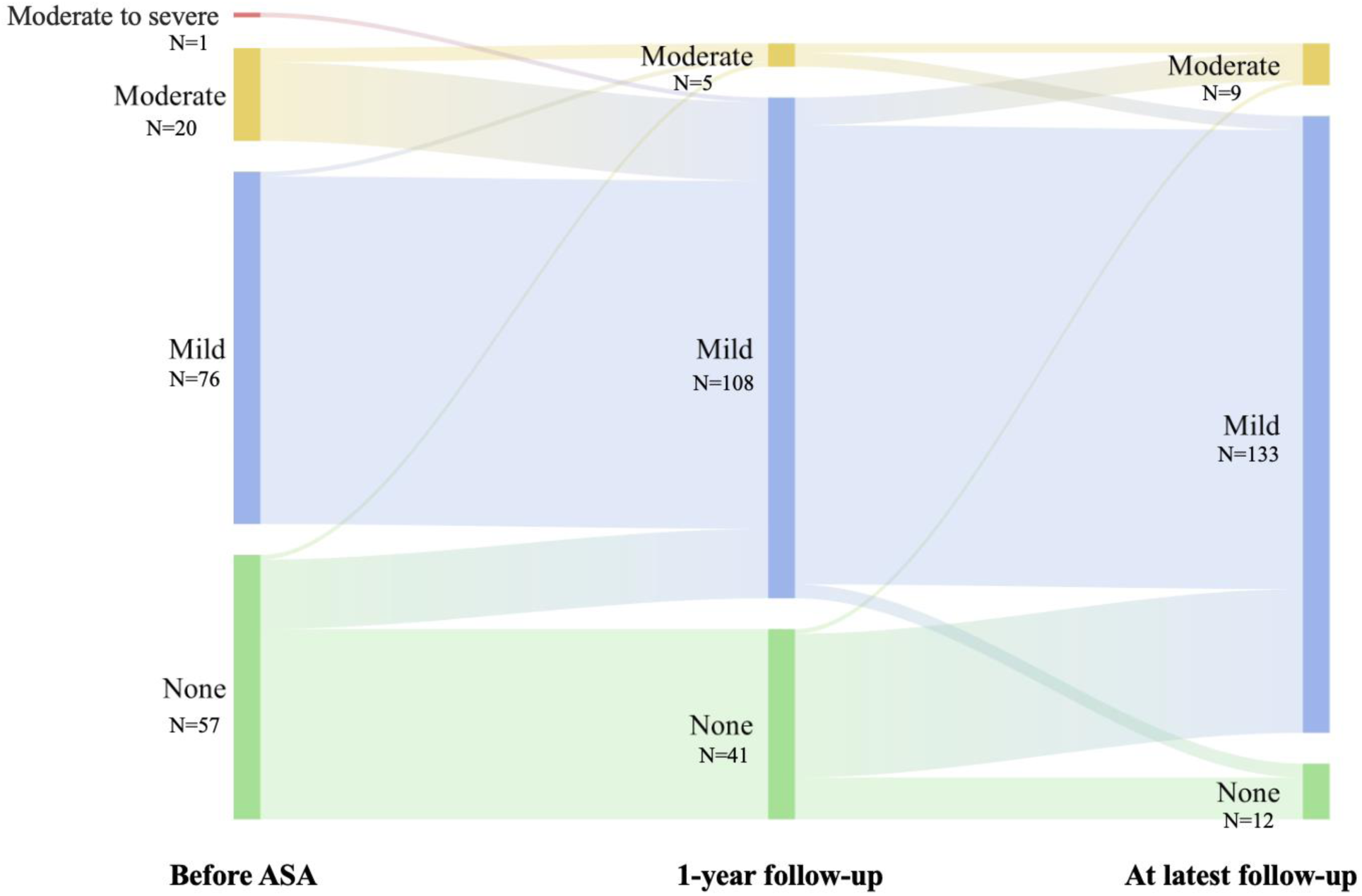
Mitral regurgitation grades at baseline, 1-year follow-up and at latest registered follow-up among patients with hypertrophic obstructive cardiomyopathy prior to, and after alcohol septal ablation. Mitral regurgitation grade (non, mild, moderate, or severe grade) at baseline before ASA (left), 1-year follow-up (middle), and latest registered follow-up (right). Abbreviations: ASA, alcohol septal ablation; MR, mitral regurgitation.

**Table 1.**
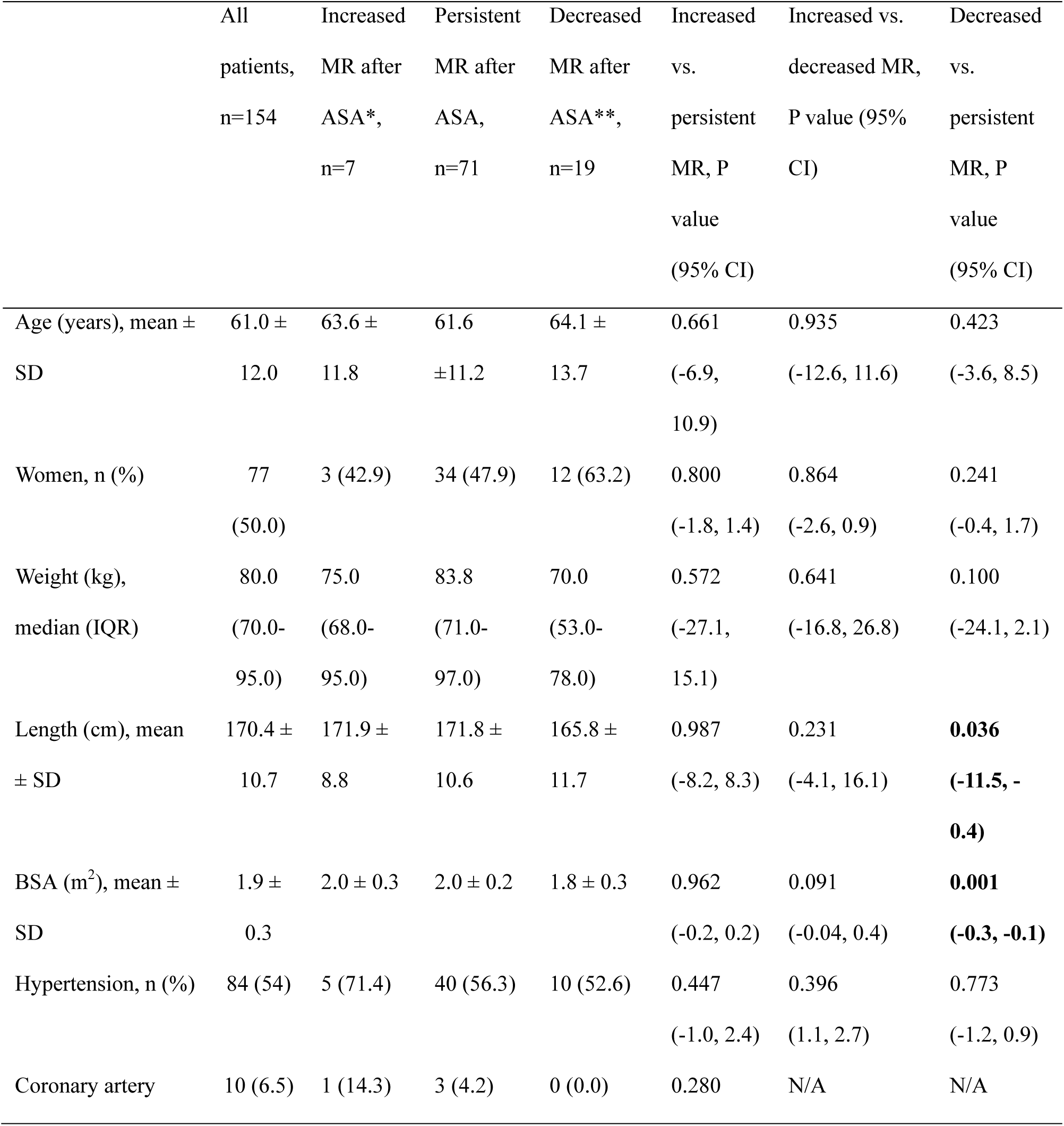
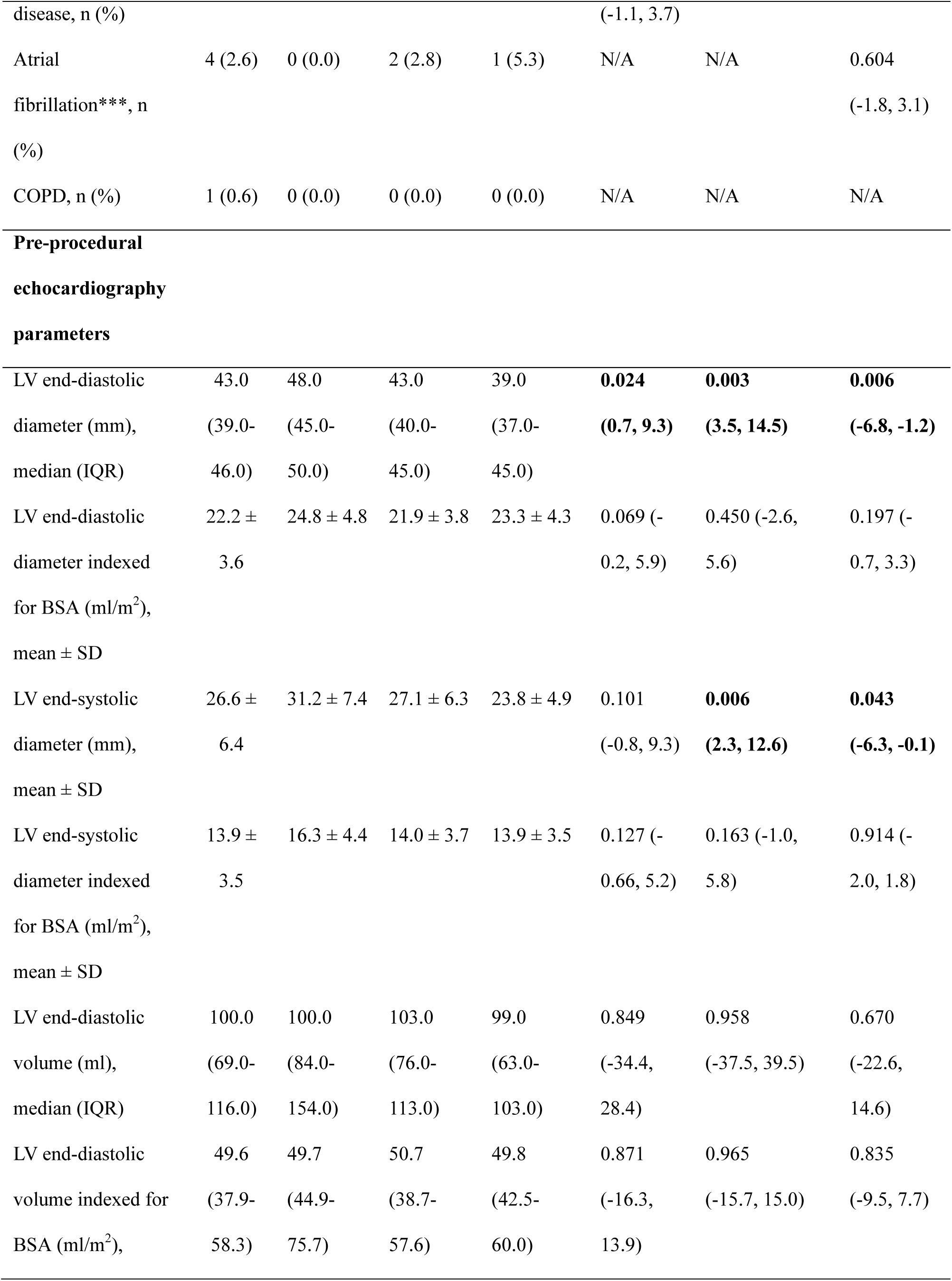
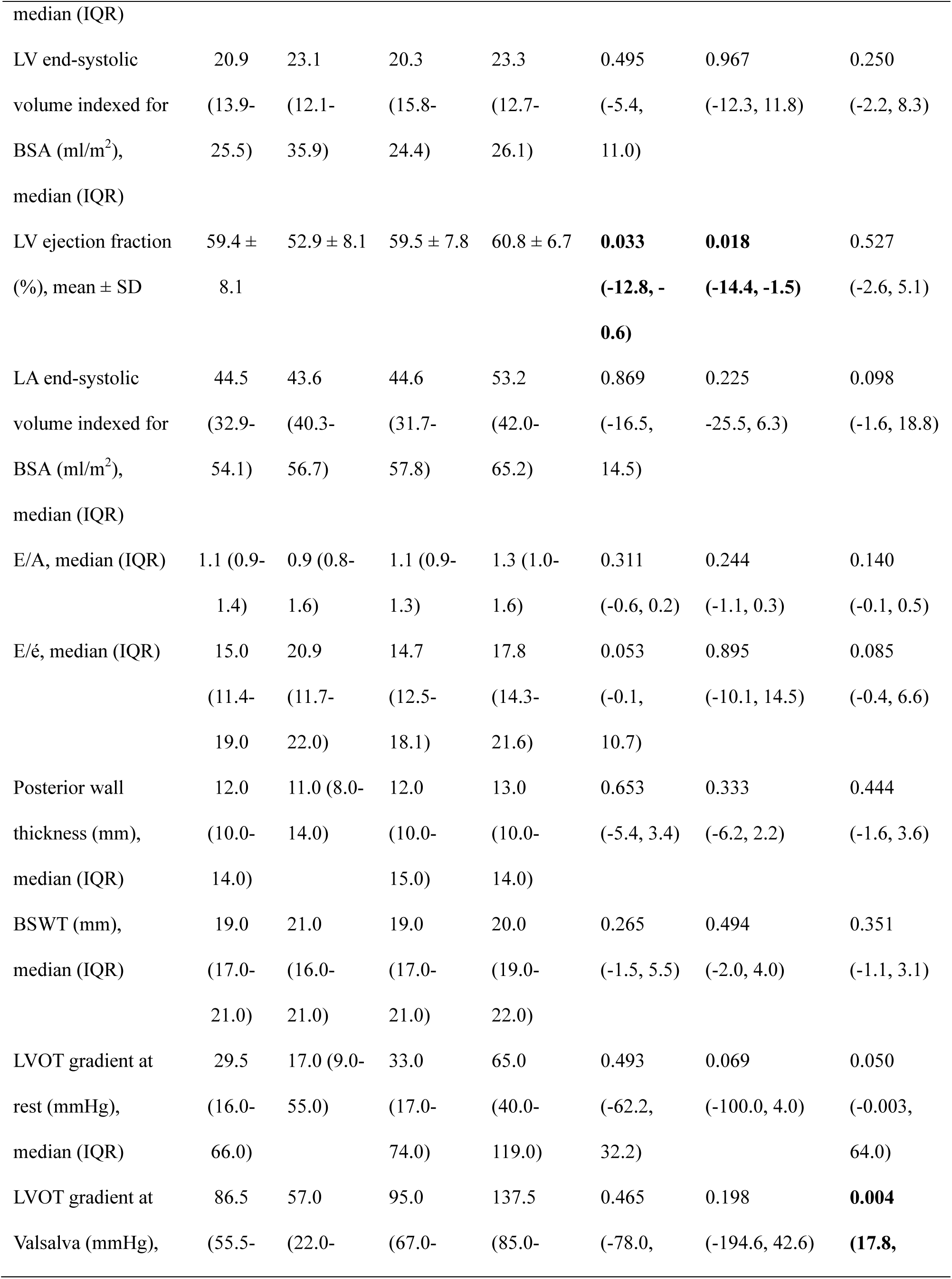
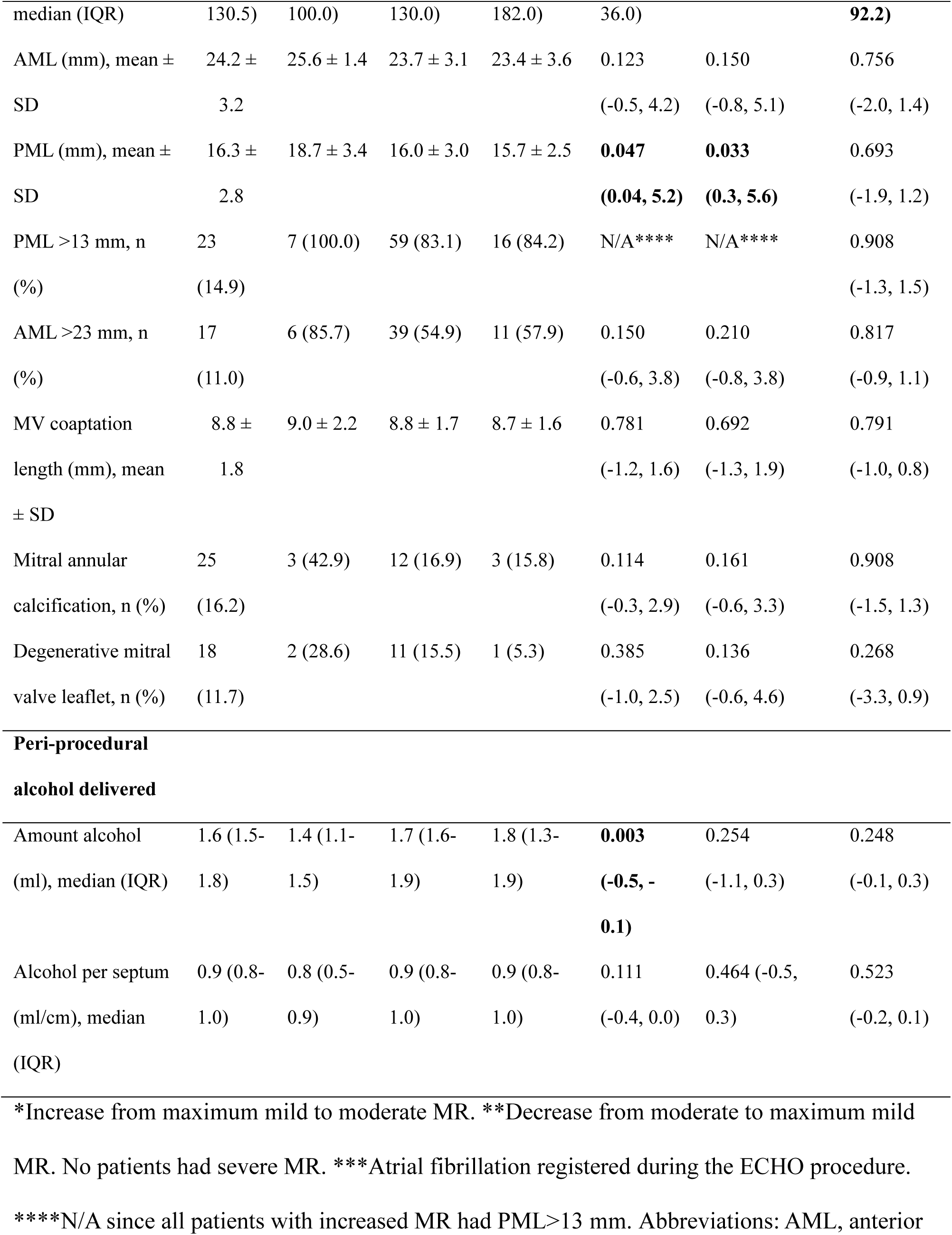
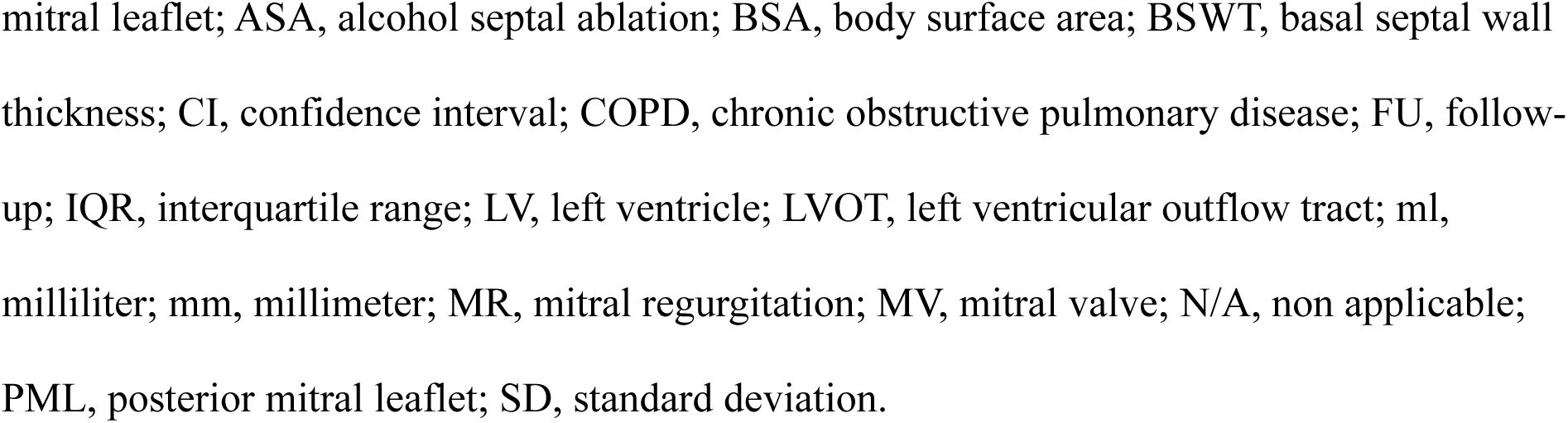
Clinical characteristics and preprocedural echocardiography parameters among patients with hypertrophic obstructive cardiomyopathy that underwent alcohol septal ablation.

**Table 2.** Mitral regurgitation severity among patients with hypertrophic obstructive cardiomyopathy before alcohol septal ablation and at follow-up.

|  | Prior to ASA, n (%). | At 1 year FU after ASA, n | At the latest FU after ASA, n |
| --- | --- | --- | --- |
|  | N=154 | (%). N=154 | (%). N=154 |
| No regurgitation | 57 (37.0) | 41 (26.7) | 12 (7.8) |
| Mild | 76 (49.4) | 108 (70.1) | 133 (86.4) |
| Moderate | 20 (13.0) | 5 (3.2) | 9 (5.8) |
| Moderate to severe | 1 (0.6) | 0 (0.0) | 0 (0.0) |
| Severe | 0 (0.0) | 0 (0.0) | 0 (0.0) |

Abbreviations: ASA, alcohol septal ablation; FU, follow-up.

There was no difference in prevalence of comorbidities among the patients who increased, had persistent grade of MR, or decreased in grade of MR (Table 1). Comparisons of ECHO parameters between the pre-procedural- and last follow-up ECHO are presented in Table 3. Three patients underwent MV replacement with mechanical mitral prosthesis after ASA, one due to mitral valve infective endocarditis, and two due to severe obstruction and SAM despite both ASA and myectomy (all had maximum mild pre-operative MR). In total 8.4% (n=13) patients underwent redo ASA during the follow-up period. One of these patients had increased MR at the last follow-up (despite redo ASA), one had decreased MR at the last follow-up, and 6 had persistent MR. Twelve of the patients underwent the redo ASA >1 year after the first ASA and one patient after 0.53 years (that patient had persistent grade of MR at both 1-year follow-up and the last follow-up). Six patients underwent surgical myectomy after ASA, 50% with no- and 50% with mild pre-procedural MR. All had persistent MR grade at follow-up except one patient who underwent myectomy 7 months after ASA and increased from no pre-procedural MR to moderate MR 4.2 years after the myectomy.

**Table 3.**
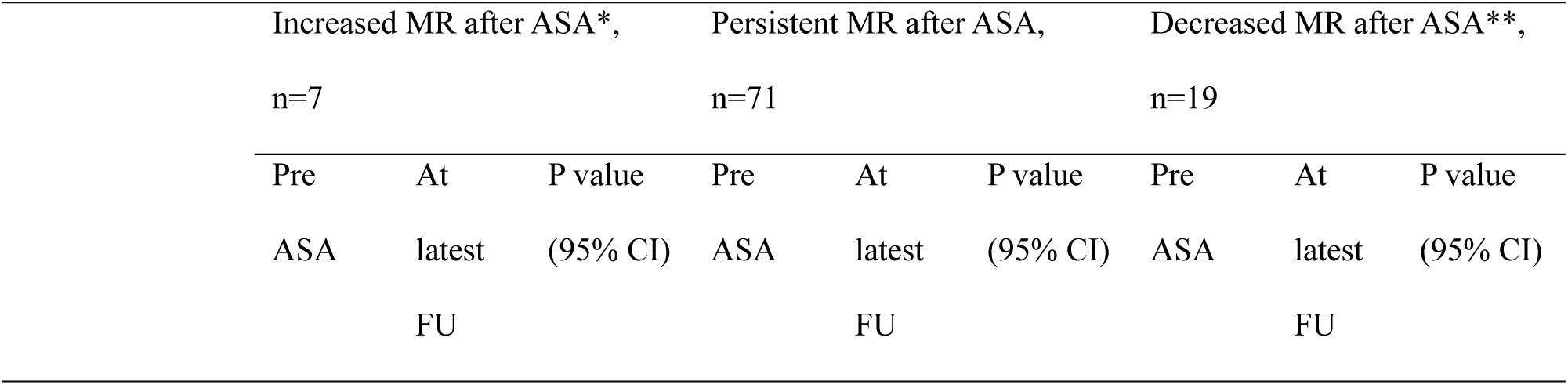
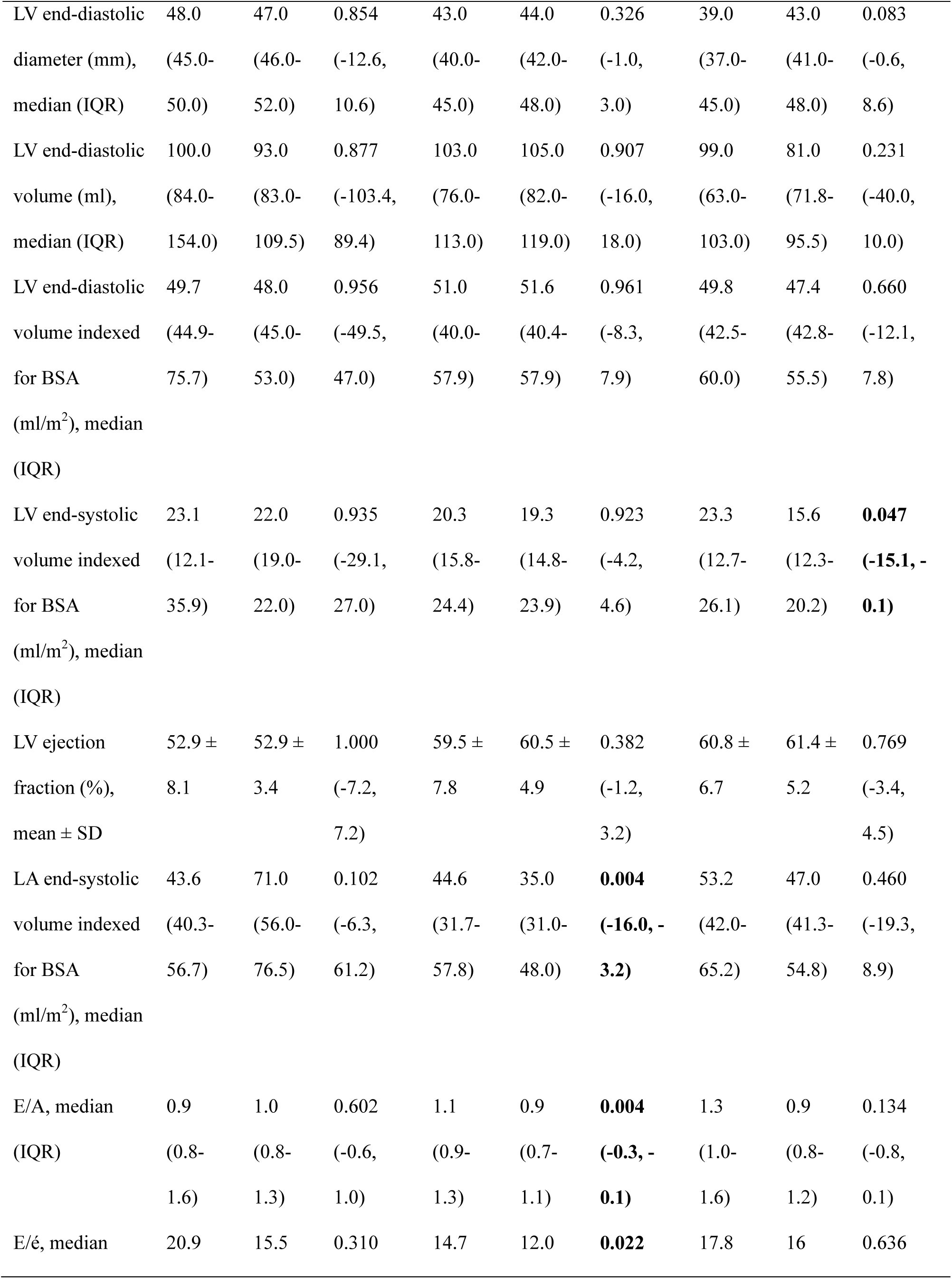
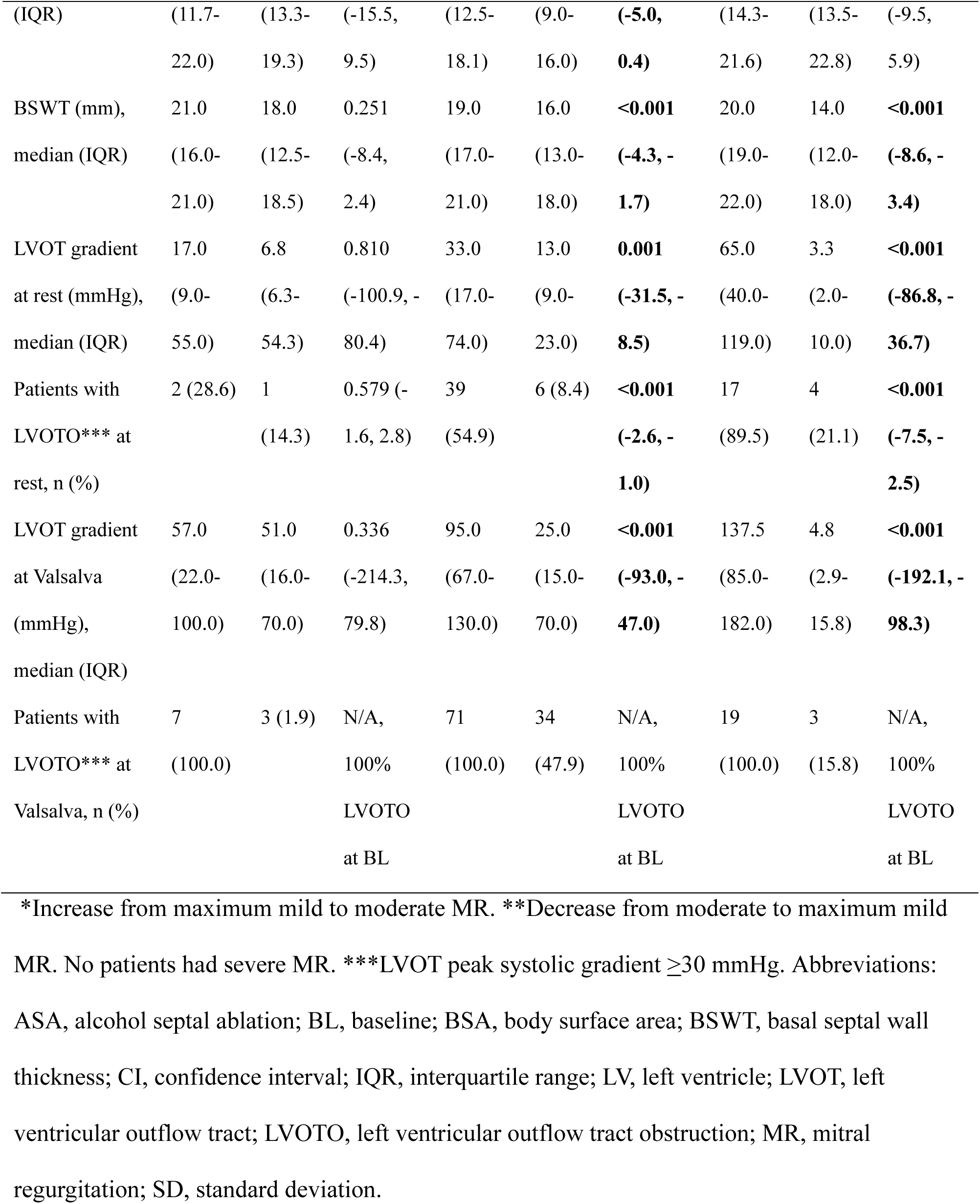
Comparison of echocardiography parameters related to mitral regurgitation among patients with hypertrophic obstructive cardiomyopathy undergoing alcohol septal ablation.

| Increased MR after ASA*,<br>n=7 |  |  | Persistent MR after ASA,<br>n=71 |  |  | Decreased MR after ASA**,<br>n=19 |  |  |
| --- | --- | --- | --- | --- | --- | --- | --- | --- |
| Pre | At | P value | Pre | At | P value | Pre | At | P value |
| ASA | latest | (95% CI) | ASA | latest | (95% CI) | ASA | latest | (95% CI) |
|  | FU |  |  | FU |  |  | FU |  |

|  |  |  |  |  |  |  |  |  |  |
| --- | --- | --- | --- | --- | --- | --- | --- | --- | --- |
| LV end-diastolic diameter (mm), median (IQR) | 48.0<br>(45.0-50.0) | 47.0<br>(46.0-52.0) | 0.854<br>(-12.6, 10.6) | 43.0<br>(40.0-45.0) | 44.0<br>(42.0-48.0) | 0.326<br>(-1.0, 3.0) | 39.0<br>(37.0-45.0) | 43.0<br>(41.0-48.0) | 0.083<br>(-0.6, 8.6) |
| LV end-diastolic volume (ml), median (IQR) | 100.0<br>(84.0-154.0) | 93.0<br>(83.0-109.5) | 0.877<br>(-103.4, 89.4) | 103.0<br>(76.0-113.0) | 105.0<br>(82.0-119.0) | 0.907<br>(-16.0, 18.0) | 99.0<br>(63.0-103.0) | 81.0<br>(71.8-95.5) | 0.231<br>(-40.0, 10.0) |
| LV end-diastolic volume indexed for BSA (ml/m<sup>2</sup>), median (IQR) | 49.7<br>(44.9-75.7) | 48.0<br>(45.0-53.0) | 0.956<br>(-49.5, 47.0) | 51.0<br>(40.0-57.9) | 51.6<br>(40.4-57.9) | 0.961<br>(-8.3, 7.9) | 49.8<br>(42.5-60.0) | 47.4<br>(42.8-55.5) | 0.660<br>(-12.1, 7.8) |
| LV end-systolic volume indexed for BSA (ml/m<sup>2</sup>), median (IQR) | 23.1<br>(12.1-35.9) | 22.0<br>(19.0-22.0) | 0.935<br>(-29.1, 27.0) | 20.3<br>(15.8-24.4) | 19.3<br>(14.8-23.9) | 0.923<br>(-4.2, 4.6) | 23.3<br>(12.7-26.1) | 15.6<br>(12.3-20.2) | **0.047**<br>**(-15.1, -0.1)** |
| LV ejection fraction (%), mean ± SD | 52.9 ± 8.1 | 52.9 ± 3.4 | 1.000<br>(-7.2, 7.2) | 59.5 ± 7.8 | 60.5 ± 4.9 | 0.382<br>(-1.2, 3.2) | 60.8 ± 6.7 | 61.4 ± 5.2 | 0.769<br>(-3.4, 4.5) |
| LA end-systolic volume indexed for BSA (ml/m<sup>2</sup>), median (IQR) | 43.6<br>(40.3-56.7) | 71.0<br>(56.0-76.5) | 0.102<br>(-6.3, 61.2) | 44.6<br>(31.7-57.8) | 35.0<br>(31.0-48.0) | **0.004**<br>**(-16.0, -3.2)** | 53.2<br>(42.0-65.2) | 47.0<br>(41.3-54.8) | 0.460<br>(-19.3, 8.9) |
| E/A, median (IQR) | 0.9<br>(0.8-1.6) | 1.0<br>(0.8-1.3) | 0.602<br>(-0.6, 1.0) | 1.1<br>(0.9-1.3) | 0.9<br>(0.7-1.1) | **0.004**<br>**(-0.3, -0.1)** | 1.3<br>(1.0-1.6) | 0.9<br>(0.8-1.2) | 0.134<br>(-0.8, 0.1) |
| E/$\dot{e}$, median | 20.9 | 15.5 | 0.310 | 14.7 | 12.0 | **0.022** | 17.8 | 16 | 0.636 |
| (IQR) | (11.7-22.0) | (13.3-19.3) | (-15.5,<br>9.5) | (12.5-18.1) | (9.0-16.0) | **(-5.0,**<br>**0.4)** | (14.3-21.6) | (13.5-22.8) | (-9.5,<br>5.9) |
| BSWT (mm), | 21.0 | 18.0 | 0.251 | 19.0 | 16.0 | **<0.001** | 20.0 | 14.0 | **<0.001** |
| median (IQR) | (16.0-21.0) | (12.5-18.5) | (-8.4,<br>2.4) | (17.0-21.0) | (13.0-18.0) | **(-4.3, -**<br>**1.7)** | (19.0-22.0) | (12.0-18.0) | **(-8.6, -**<br>**3.4)** |
| LVOT gradient | 17.0 | 6.8 | 0.810 | 33.0 | 13.0 | **0.001** | 65.0 | 3.3 | **<0.001** |
| at rest (mmHg), | (9.0-55.0) | (6.3-54.3) | (-100.9, -<br>80.4) | (17.0-74.0) | (9.0-23.0) | **(-31.5, -**<br>**8.5)** | (40.0-119.0) | (2.0-10.0) | **(-86.8, -**<br>**36.7)** |
| Patients with | 2 (28.6) | 1 | 0.579 (- | 39 | 6 (8.4) | **<0.001** | 17 | 4 | **<0.001** |
| LVOTO*** at |  | (14.3) | 1.6, 2.8) | (54.9) |  | **(-2.6, -**<br>**1.0)** | (89.5) | (21.1) | **(-7.5, -**<br>**2.5)** |
| rest, n (%) |  |  |  |  |  |  |  |  |  |
| LVOT gradient | 57.0 | 51.0 | 0.336 | 95.0 | 25.0 | **<0.001** | 137.5 | 4.8 | **<0.001** |
| at Valsalva | (22.0-100.0) | (16.0-70.0) | (-214.3,<br>79.8) | (67.0-130.0) | (15.0-70.0) | **(-93.0, -**<br>**47.0)** | (85.0-182.0) | (2.9-15.8) | **(-192.1, -**<br>**98.3)** |
| (mmHg), |  |  |  |  |  |  |  |  |  |
| median (IQR) |  |  |  |  |  |  |  |  |  |
| Patients with | 7 | 3 (1.9) | N/A, | 71 | 34 | N/A, | 19 | 3 | N/A, |
| LVOTO*** at | (100.0) |  | 100% | (100.0) | (47.9) | 100% | (100.0) | (15.8) | 100% |
| Valsalva, n (%) |  |  | LVOTO |  |  | LVOTO |  |  | LVOTO |
|  |  |  | at BL |  |  | at BL |  |  | at BL |

\*Increase from maximum mild to moderate MR. \*\*Decrease from moderate to maximum mild
MR. No patients had severe MR. \*\*\*LVOT peak systolic gradient $\geq 30$ mmHg. Abbreviations:
ASA, alcohol septal ablation; BL, baseline; BSA, body surface area; BSWT, basal septal wall thickness; CI, confidence interval; IQR, interquartile range; LV, left ventricle; LVOT, left ventricular outflow tract; LVOTO, left ventricular outflow tract obstruction; MR, mitral regurgitation; SD, standard deviation.

**Table 4.** Mitral valve morphology in patients with hypertrophic obstructive cardiomyopathy at the latest follow-up after alcohol septal ablation.

|  | All<br>patients,<br>n=154 | Increased<br>MR after<br>ASA, n=7 | Persistent<br>grade of<br>MR, n=71 | Decreased<br>MR after<br>ASA, n=19 | Increased<br>vs.<br>persistent<br>MR, P value<br>(95% CI) | Increased<br>vs.<br>decreased<br>MR, P value<br>(95% CI) | Decreased<br>vs. persistent<br>MR, P value<br>(95% CI) |
| --- | --- | --- | --- | --- | --- | --- | --- |
| Mitral annular<br>calcification, n<br>(%) | 37 (24.0) | 4 (57.1) | 18 (25.4) | 5 (26.3) | 0.092<br>(-0.2, 3.0) | 0.143<br>(-0.5, 3.1) | 0.932<br>(-1.1, 1.2) |
| Degenerative<br>mitral valve<br>leaflet, n (%) | 28 (18.2) | 4 (57.1) | 12 (16.9) | 2 (10.5) | **0.005**<br>**(0.8, 4.3)** | **0.006**<br>**(0.9, 5.3)** | 0.500<br>(-2.1, 1.0) |

Abbreviations: AML, anterior mitral leaflet; ASA, alcohol septal ablation; CI, confidence interval; MR, mitral regurgitation; PML, posterior mitral leaflet.

All 21 patients that had at least moderate MR prior to ASA were evaluated as primarily SAM-mediated etiology at baseline. Degenerative abnormalities such as mitral annular calcification and leaflet degeneration were rare at baseline, whereas elongated mitral valve leaflets (Table 1) were more common, potential contributors to SAM. No patient presented with MV prolapse, chordal rupture, leaflet destruction, or rheumatic disease, although one patient had an anomalous papillary muscle attachment, and four patients including the one with anomalous papillary muscle attachment, had exceptionally elongated chordae tendinea. At the 1-year follow-up, SAM was reduced in 85.7% of the 21 patients with at least moderate MR prior to ASA resulting in reduction to mild grade of MR, three had persistent SAM and moderate MR at 1-year follow-up but reduced to mild MR at the last follow-up related to reduced SAM. At the last follow-up, two of the 18 patients who improved to mild MR at 1-year follow-up had progressed in basal septal wall thickness with worsened SAM and LVOTO, resulting in moderate grade of MR again at the last follow-up (Fig 1). That resulted in 90.5% of the 21 patients with at least moderate MR prior to ASA reduced to mild MR at the last follow-up. The patients with decreased or persistent grade of MR showed significant reduction of basal septal wall thickness and LVOT pressure gradients at follow-up, in contrary to the patients with increased MR grade (Table 3). When comparing the absolute reduction in basal septal wall thickness between the groups, patients with increased MR grade tended to exhibit a smaller reduction than those with decreased MR grade, although the difference was not statistically significant (Table S1). Similarly, patients with an increased MR grade demonstrated a smaller reduction in LVOT gradients during the Valsalva maneuver than patients with a decreased MR grade, but this trend also did not reach statistical significance (Table S1). Further, patients with increased MR grade tended to have a smaller reduction of LVOT gradients during Valsalvas maneuver, compared with the group of patients with decreased MR grade, however not statistically significant (Table S1). The amount of alcohol delivered did not affect the outcome in regard to MR (Table 1).

Of the seven patients with increased MR grade, the mechanisms of MR were increased severity of SAM (n=3), degenerative etiology that was not causing a MR at the baseline ECHO nor at the one-year follow-up (n=3, revealed at last follow-up ECHO 3.8 years, 4.8 years and 5.2 years respectively after the procedure), and one secondary MR due to enlarged atrium revealed at the one-year follow-up. Degenerative mitral valve disease at follow-up and elongated posterior mitral valve leaflet were more common among the patients that increased the grade of MR, compared both with patients with decreased grade of MR and those with persistent grade of MR at follow-up, respectively (Table 1 and 4).

Preprocedural LV ejection fraction was lower among the patients who had increased MR at follow-up, compared with those who had decreased or persistent grade of MR at follow-up (Table 1 and 5). When using a cutoff for enlarged LV end-diastolic (>50 mm or >45 mm for men and women respectively), LV ejection fraction <55%, and elongated PML >13 mm (21, 23), patients with increased MR grade at follow-up fulfilled these parameters in a higher extent compared with patients with decreased or persistent MR grade at follow-up (Table 5). Those who increased their MR grade after ASA had an increase (although not significant) in left atrial volume, but those who decreased in MR had a slight (but not significant) decrease in left atrial volume (Table 3). No significant abnormalities were seen in the aortic-, tricuspid-, or pulmonary valves.

**Table 5.** Comparison of pre-procedural echocardiography parameters among patients with hypertrophic obstructive cardiomyopathy having either increase or decrease of mitral regurgitation at follow-up after alcohol septal ablation.

|  | All<br>patients,<br>n=154 | Increased<br>MR after<br>ASA, n=7 | Persistent<br>grade of<br>MR, n=71 | Decreased<br>MR after<br>ASA, n=19 | Increased<br>vs.<br>persistent<br>MR, P value<br>(95% CI) | Increased vs.<br>decreased<br>MR, P value<br>(95% CI) | Decreased<br>vs. persistent<br>MR, P value<br>(95% CI) |
| --- | --- | --- | --- | --- | --- | --- | --- |
| Enlarged<br>LVEDVi*, n | 74 (48.1) | 4 (57.1) | 37 (52.1) | 8 (5.2) | 0.800<br>(-1.4, 1.8) | 0.498<br>(-1.1, 2.4) | 0.440<br>(-1.4, 0.6) |
| (%) |  |  |  |  |  |  |  |
| Enlarged | 21 (13.6) | 4 (57.1) | 9 (12.7) | 2 (10.5) | **0.009** | **0.023** | 0.800 |
| LVEDd**, n |  |  |  |  | **(0.6, 3.9)** | **(0.3, 4.5)** | (-1.8, 1.4) |
| (%) |  |  |  |  |  |  |  |
| LVEF<55%, | 27 (22.5) | 4 (57.1) | 10 (14.1) | 1 (5.3) | **0.012** | **0.013** | 0.318 |
| n (%) |  |  |  |  | **(0.5, 3.7)** | **(0.7, 5.7)** | (-3.2, 1.0) |

\* LVEDVi >54 resp 45 ml/m<sup>2</sup> for men and women respectively. \*\*LVEDd >50 resp 45 mm for men resp women. Abbreviations: ASA, alcohol septal ablation; CI, confidence interval; LVEDd, left ventricular end-diastolic diameter; LVEDVi, left ventricular end-diastolic volume indexed for body surface area; LVEF, left ventricular ejection fraction; MR, mitral regurgitation.

## Discussion

With a mean follow-up time after ASA of 4.0 ± 3.9 years we observed an overall favorable trend, with most patients demonstrating a decrease in MR grade after ASA. However, a subset of patients showed the opposite pattern, developing moderate MR after ASA. This suggests that septal reduction therapy was either insufficient to alleviate SAM-related MR, or that patients developed additional LV and/or MV pathology contributing to MR progression. Prior studies have reported diverging effects on MR after surgical septal myectomy: with some showing marked improvement in SAM-mediated MR, and others with residual or progressive MR over time (13, 24, 25). To our knowledge, this study is among the few that specifically examine the outcome of MR after ASA, and includes one of the largest patient cohorts to date (15–17). Although previous studies of ASA did not designate MR as a primary outcome or assess ECHO predictors of MR progression, all studies showed reduced rates of moderate to severe MR after ASA (15–17). A Chinese study of 46 HOCM patients undergoing ASA demonstrated a reduction from 79.1% (n=34) with moderate to severe MR at baseline to 13.9% (n=6) at 14 months follow-up ECHO (15). A Portuguese study of 80 patients with mean follow-up 4.17±2.13 years showed a reduction from 32.5% (n=26) moderate MR at baseline to 16.3% (n=13) at the 14 months follow-up ECHO (16). A Japanese study of 44 patients with median 6 years follow-up showed a reduction from 53% (n=23) moderate or severe MR at baseline to 16% (n=7) at follow-up (17). These reductions are broadly consistent with our findings, which showed a decrease from 13.6% (n = 21) with moderate or moderate-to-severe MR at baseline to 5.8% (n = 9) at last follow-up, although our cohort had a lower baseline prevalence of moderate MR.

SAM-mediated MR is the most common MR mechanism in HOCM (25). Adequate surgical septal reduction therapy via myectomy has been shown to significantly decrease SAM-related MR (13, 24, 25, 26). The most favorable outcomes after isolated myectomy have been observed in patients with marked septal hypertrophy, provided the procedure sufficiently enlarges the LVOT (13, 24–26). In short, the greater the degree of septal hypertrophy, the lower the likelihood that concomitant MV intervention is required (26). In our study, most patients with moderate MR prior to ASA demonstrated improvement in MR grade at follow-up. These patients showed significant reductions in basal septal wall thickness, LVOT pressure gradients, and SAM, likely reflecting LVOT enlargement achieved through ASA.

Among the seven patients who developed moderate MR after ASA, three exhibited worsening SAM, three had degenerative MR (one detected at 1-year follow-up and two at final follow-up), and one developed MR secondary to left atrial enlargement. Degenerative leaflet changes and elongated posterior mitral leaflets at baseline were more prevalent among patients who increased in MR grade, compared with those with decreased or persistent grade of MR at follow-up.

Three patients underwent MV replacement with a mechanical prosthesis after ASA: one due to infective endocarditis and two due to persistent severe LVOT obstruction and SAM despite both ASA and subsequent myectomy. Notably, all three had no or only mild MR prior to ASA at and follow-up. Among the patients who increased in MR grade after ASA, several echocardiographic factors beyond basal septal wall thickness may have contributed to the worsening of MR. Lower LV ejection fraction, longer posterior mitral leaflet length, and higher prevalence of degenerative mitral valve leaflets were observed in this subgroup. Enlarged left atrial volume and reduced systolic LV function may predispose to functional MR, whereas elongated posterior leaflets may promote MR secondary to SAM or impaired MV coaptation. Importantly, these factors would not be addressed by septal reduction therapy alone. Multiple studies highlight the need for concomitant MV interventions in patients with non-SAM-dependent MR (13, 25, 26). Identifying intrinsic MV disease and other contributors to MR prior to septal reduction therapy in HOCM is therefore important, particularly in patients with mild septal hypertrophy who may derive greater benefit from MV repair (27, 28). Patel et al., demonstrated that patients with basal septal thickness ≤1.8 cm and MV anomalies frequently required concomitant MV procedures (28).

### Limitations

One limitation of this study is the single-center design, possibly limiting the generalizability. However, because this center is a tertiary referral institution receiving patients from different regions of Sweden, the study population effectively reflects a national cohort, enhancing generalizability relative to a typical single-center study. Although this is one of the largest ASA centers in Sweden, the absolute number of events (i.e., MR of at least moderate grade) was low, limiting power for multivariable analyses and necessitating reliance on univariate methods.

All ASA referrals were evaluated by our multidisciplinary team and decisions on septal reduction therapy adhered to contemporary European guidelines (14), limiting selection bias. Potential information bias related to retrospective echocardiographic review, including MR grading, was mitigated by conducting all assessments independently of clinical reports. After all echocardiograms were reviewed, any discrepancies between study and clinical reports were resolved through consensus with at least one additional cardiovascular imaging specialist, thereby minimizing misclassification, observer, and reporting bias.

### Future directions

The results from this study suggest that ECHO evaluation of the left ventricle and mitral valve prior to ASA may predict the worsening or development of MR after ASA. However, the results could benefit from future validation in a prospective, multicenter approach with a larger study population and longer follow-up. A longer follow-up would possibly detect even further degenerative MV lesions; and possibly further clinical outcomes such as heart failure events, quality of life, and mortality triggered by MR after ASA.

## Conclusion

In our retrospective study of HOCM patients undergoing ASA, most patients with at least moderate MR prior to ASA regressed to mild or less MR at follow-up, likely reflecting LVOT enlargement and reduction of SAM following septal reduction. However, a subset of patients exhibited increased grade of MR, in whom intrinsic MV disease such as degenerative, or elongated mitral leaflets, along with depressed LV function, and enlarged left atrium, may have contributed to MR progression. Although studies with larger sample sizes are needed to confirm these findings, our results suggest that specific pre-procedural echocardiographic features may help identify patients at increased risk of MR deterioration after ASA, thereby improving risk stratification and informing decisions regarding concomitant MV repair or replacement.

## Sources of funding

A.D. would like to acknowledge funding from the Swedish Heart-Lung foundation (project number 20230742). D.M. would like to acknowledge funding from the Swedish Heart-Lung foundation (project number 20230080), and the European Union (ERC, MultiPRESS, 101075494). Views and opinions expressed are those of the authors and do not reflect those of the Swedish Heart-Lung foundation, the European Union, or the European Research Council Executive Agency.

## Disclosures

Nothing to report

## Ethical approval

This study was performed in line with the principles of the Declaration of Helsinki. Approval was granted by the Swedish Ethical Review Authority with the registration number: 2022-01472-01 and 2024-04242-01.

## Consent to participate

This study does not present any identifying details. All information was anonymized, and the study did not imply any risks for the patients since it did not affect the patients or cause any changes in their treatments or care.

## Author contributions

The study was designed by AD. The material was prepared by AD, AR, MJE, ARü, and NS. Data collection was performed by AD, MM, and RJ. Data analysis was performed by AD. AD wrote the first draft of this manuscript with a first review by ARü and DM, and all authors commented on the following versions of the manuscript. All authors have read and approved the final manuscript.

## Data Availability

Consideration upon request.

## Abbreviations

ASA: alcohol septal ablation
ECHO: echocardiography
HOCM: hypertrophic obstructive cardiomyopathy
LV: left ventricle
LVOT: left ventricular outflow tract
LVOTO: left ventricular outflow tract obstruction
MR: mitral regurgitation
MV: mitral valve
SAM: systolic anterior motion

## Supplemental material

**Table S1.** Differences in left ventricular outflow tract peak systolic gradient and basal septal wall thickness in patients with hypertrophic obstructive cardiomyopathy after alcohol septal ablation.

|  | All patients, n=154 | Increased MR after ASA, n=7 | Persistent grade of MR, n=71 | Decreased MR after ASA, n=19 | Increased vs. persistent MR, P value (95% CI) | Increased vs. decreased MR, P value (95% CI) | Decreased vs. persistent MR, P value (95% CI) |
| --- | --- | --- | --- | --- | --- | --- | --- |
| Difference in basal septal wall thickness (mm), mean ± SD | -3.4 ± 4.0 | -1.5 ± 4.2 | 3.5 ± 4.1 | -4.6 ± 4.1 | 0.267 (-1.5, 5.5) | 0.120 (-0.9, 7.1) | 0.281 (-3.3, 1.0) |
| Difference in LVOT gradient at rest (mmHg), median (IQR) | -12.0 (-48.0- -3.0) | -1.5 (-46.0-0.0) | -12.0 (-50.0- -4.0) | -7 (-24.0- -2.0) | 0.666 (-32.3, 50.3) | 0.205 (-23.4, 103.4) | 0.050 (-61.9, -0.1) |
| Difference in LVOT gradient at Valsalva maneuver (mmHg), median (IQR) | -48.0 (-90.0- -11.0) | -2.0 (-90.0-7.0) | -61.0 (-90.0- -20.0) | -87.5 (-130.0- -28.0) | 0.095 (-10.6, 128.6) | 0.146 (-30.7, 192.7) | 0.330 (-66.7, 22.7) |

Measures include differences between baseline ECHO and the latest follow-up ECHO registered.
Abbreviations: ASA, alcohol septal ablation; CI, confidence interval; IQR, interquartile range; mmHg, millimeters of Mercury; MR, mitral regurgitation; SD, standard deviation.

## Notes

### Competing Interest Statement

The authors have declared no competing interest.

### Clinical Trial

Retrospective study

